# Determinants of Public Participation in Primary Health Care Worldwide: A Scoping Review Protocol

**DOI:** 10.1101/2024.12.11.24318663

**Authors:** Kimia Jazi, Amirhosein Mahmoodi, Mohammadreza Azizkhani, hassan joulaei

## Abstract

Public participation is a key component to achieve effective primary health care (PHC) and considering human rights to health. Multiple factors affect community involvement regarding varied health systems worldwide. Obstacles should be addressed in each country, regarding health system regulations and community willingness. This scoping review will systematically map literature on varied models, potential barriers, as well as challenges and future opportunities to public participation in PHC around the world.

This study will follow and will be conducted through Joanna Briggs Institute (JBI) guidelines. The PRISMA Extension for Scoping Reviews (PRISMA-ScR) will also guide the reporting. The search will be conducted across PubMed, Scopus, Web of Science Core Collection, Cochrane Central Register of Controlled Trials Embase, SID, and Magiran databases focusing on studies published from 2010 to January 2025 in English and Farsi. Two independent reviewers will screen titles and abstracts, followed by full-text screening. Data will be extracted in a standardized form by two authors and all disparities will be resolved through consulting a third author. Diagrams, tables, and figures will be applied to illustrate extracted data.

## Introduction

### Background

Public participation in primary health care (PHC) was first considered in Alma-Ata Declaration 1978, highlighting the right of involving in healthcare planning as a significant effective and controlling factor (1). Similarly, the World Health Organization (WHO) defines community participation as an active involvement in making decisions on what affects their lives and associated policies (2, 3). In societies where governments fail to provide adequate public health services, social involvement plays a crucial role in prioritizing global health issues (4). Besides resource limitations, on one hand, the health systems are exposed to many unexpected disasters worldwide. On the other hand, they face reforms every year in line with different disease patterns (5). Thus achieving “health for all” require cooperation of different organisms under participatory approaches in order to strengthen resilience and empower the society (6, 7).

Multiple studies established experiences of public and health professional involvements, confirming the positive outcomes, and underlying potential barriers (8-16). Accordingly, incompetent health workers, inconsistent community needs and education, organizational issues such as bureaucracy, structural disturbances including long-distance, inadequate participatory groups, conflict of interests, and contradictory cultural differences (14, 17-20). A more recent qualitative study in Iran categorized obstacles to community involvement in PHC in five groups including trust in health system, community and health system perception of participation, cultural, community participatory programs, and institutional and bureaucratic problems (16). Abdel Salam et al. reported that illiteracy, lower health awareness, gender disparities, and lack of participating desire significantly affect public participation in PHC in Saudi Arabia (21). Another qualitative study on Sub-Saharan population also showed that health care workers overlooked community-level health committees regarding inadequate relevant education (22).

Although various studies assessed obstacles to public participation in primary health care, as a fundamental element in patient-centered care, could be influenced by different factors such as geographical location, population, type of care, culture, education, health system regulations, and even definition of participation per se. Moreover, to our knowledge, there is no study investigating different participation barriers in varied countries worldwide. Therefore, the present scoping review aims to comprehensively investigate different models of public involvement, obstacles to community participation, and providing insights to imply the best model in primary health care around the world regarding different healthcare models and further influencing factors.

### Objectives

Current scoping review is aimed to investigate the body of literature on determinants of public participation in primary healthcare (PHC) worldwide. In particular, the research will address the following questions:

1. What are different types of public participation in PHC?
2. What are the key determinants of public participation in PHC?
3. How do these determinants influence and interact each other?
4. What are challenges and opportunities regarding key determinants and different forms of participation?
5. What are current research gaps in community participation in PHC?

Considering above questions, the aim of this study is to provide a comprehensive review of community participation in PHC and influencing factors, as well as, identifying evidence-based directions for future research and interventions.

## Methods

This scoping review will be subsequent to the methodology developed by Arksey and O’Malley (2005), and enhancements by Levac et al. (2010) (23, 24). The review protocol and the scoping review, both follows the Joanna Briggs Institute (JBI) guidelines for developing a scoping review (25). To ensure the quality and transparency of comprehensive reporting, the manuscript will be developed upon the PRISMA Extension for Scoping Reviews (PRISMA-ScR) checklist (26).

### Eligibility Criteria

- **Inclusion Criteria** Population: People of all ages. Concept: Determinants of public participation in PHC Context: PHC setting worldwide Outcomes: community participation Study Types: Original studies including experimental, cohort, cross-sectional, case-control, qualitative studies, and all types of reviews (for reference check). Language: Articles published in English and Farsi. Publication Date: Studies published from January 2010 up to January 2025.
- **Exclusion Criteria** Population: -- Intervention: Studies that have not investigated factors influencing public engagement in PHC Outcomes: Studies that do not evaluate public participation. Study Types: case reports, case series, letters, commentaries and editorials Language: Articles published in languages other than English and Farsi.
- **The rationale for Criteria:** To achieve the best outcome of PHC in the community, the engagement of population is necessary. Investigating different forms of participation, the factors determining the community participation, and comparing their role would provide promising evidence on opportunities and challenges faced PHC performance worldwide. Limiting the review to articles in English and Farsi, in addition to limiting the time of publication for the past 14 years ensures the inclusion of relevant, accessible, and up to date studies.
- **Limitations/Restrictions:** Language bias by including only Farsi and English literature.
- **Handling Ambiguous Information:** The ambiguous information will be discussed in the research group. If consensus is unattainable, a third reviewer will attend for further evaluation.
- **Shared Interpretation of Criteria:** Meeting will be held to make sure all members come into a consistent understanding of the eligibility criteria, regularly.

### Information Sources and Search Strategy

- **Databases:** PubMed, Scopus, Web of Science Core Collection, Cochrane Central Register of Controlled Trials (CENTRAL), Embase, SID, and Magiran
- **General Search Terms: ‘**Determinant*’, ‘Factor*’, ‘Model*’, ‘User involvement’, ‘Community engagement’, ‘patient involvement’, ‘primary health care’, ‘Primary Health Care’ (Table 1).
- **Citation Management Software:** EndNote will be applied for citations.
- **PRESS Checklist:** The Peer Review of Electronic Search Strategies (PRESS) checklist will be used to guarantee the quality, transparency and comprehensiveness of the search strategy.

**Table 1.**
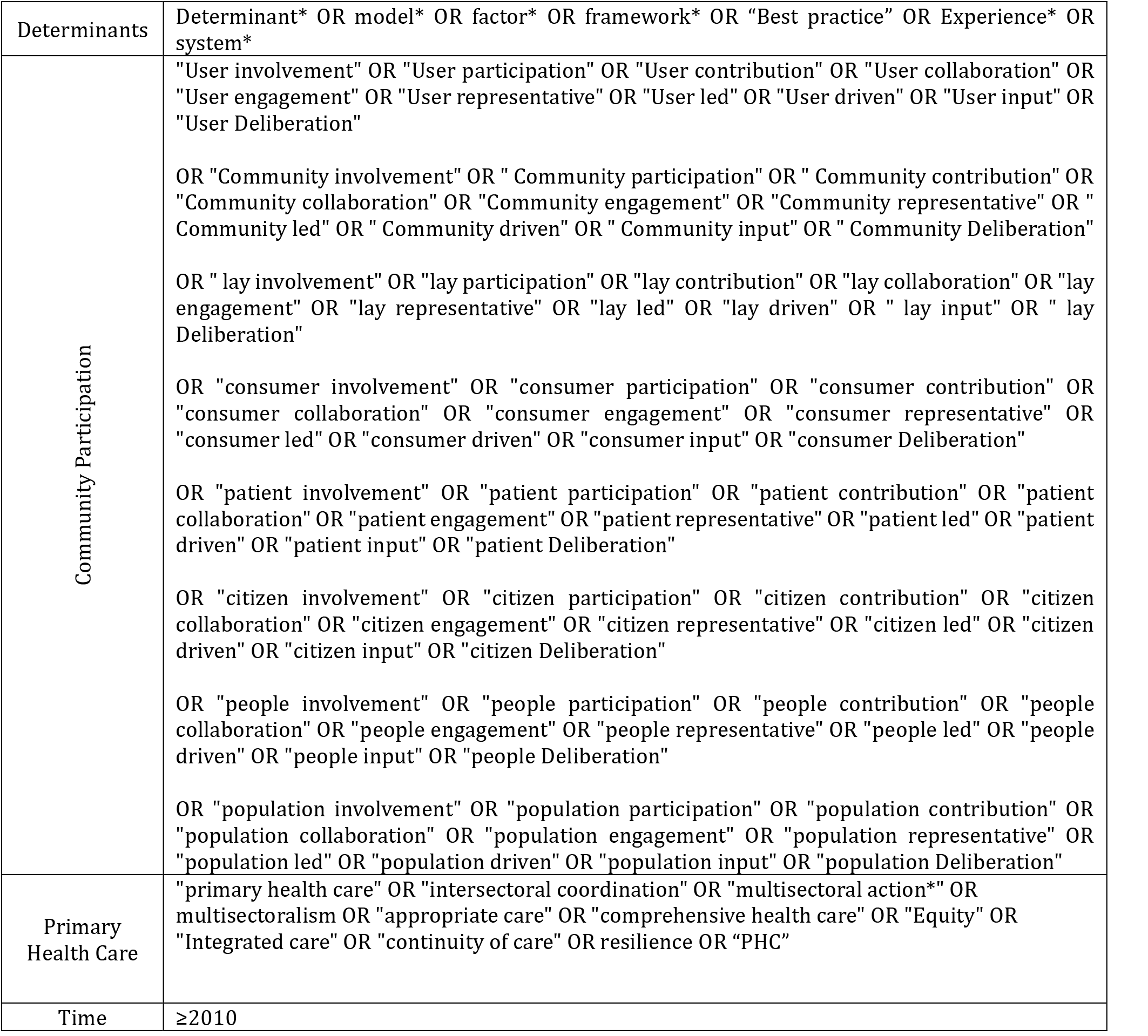

|  |  |
| --- | --- |
| Determinants | Determinant* OR model* OR factor* OR framework* OR “Best practice” OR Experience* OR system* |
| Community Participation | "User involvement" OR "User participation" OR "User contribution" OR "User collaboration" OR "User engagement" OR "User representative" OR "User led" OR "User driven" OR "User input" OR "User Deliberation"<br>OR "Community involvement" OR "Community participation" OR "Community contribution" OR "Community collaboration" OR "Community engagement" OR "Community representative" OR "Community led" OR "Community driven" OR "Community input" OR "Community Deliberation"<br>OR "lay involvement" OR "lay participation" OR "lay contribution" OR "lay collaboration" OR "lay engagement" OR "lay representative" OR "lay led" OR "lay driven" OR "lay input" OR "lay Deliberation"<br>OR "consumer involvement" OR "consumer participation" OR "consumer contribution" OR "consumer collaboration" OR "consumer engagement" OR "consumer representative" OR "consumer led" OR "consumer driven" OR "consumer input" OR "consumer Deliberation"<br>OR "patient involvement" OR "patient participation" OR "patient contribution" OR "patient collaboration" OR "patient engagement" OR "patient representative" OR "patient led" OR "patient driven" OR "patient input" OR "patient Deliberation"<br>OR "citizen involvement" OR "citizen participation" OR "citizen contribution" OR "citizen collaboration" OR "citizen engagement" OR "citizen representative" OR "citizen led" OR "citizen driven" OR "citizen input" OR "citizen Deliberation"<br>OR "people involvement" OR "people participation" OR "people contribution" OR "people collaboration" OR "people engagement" OR "people representative" OR "people led" OR "people driven" OR "people input" OR "people Deliberation"<br>OR "population involvement" OR "population participation" OR "population contribution" OR "population collaboration" OR "population engagement" OR "population representative" OR "population led" OR "population driven" OR "population input" OR "population Deliberation" |
| Primary Health Care | "primary health care" OR "intersectoral coordination" OR "multisectoral action*" OR "multisectoralism" OR "appropriate care" OR "comprehensive health care" OR "Equity" OR "Integrated care" OR "continuity of care" OR resilience OR “PHC” |
| Time | ≥2010 |

### Study Selection/Screening

- **Eligibility Criteria for Screening:** As mentioned.
- **Title and Abstract Screening:** The tile and abstract of all searched studies will be screened by two independent reviewers. In the next step, a pilot screening would be performed to ensure the consistency. Rayyan research assist will be used to manage and screen articles. Any discrepancies would be first discussed by reviewers, and if no agreement achieved, the consultation of a third reviewer would be necessary.
- **Full-Text Screening:** The articles that were found potentially relevant to the aim of the study by screening title and abstract, will be evaluated for their full-text by two reviewers independently. As mentioned, inconsistencies would be resolved by the decision of the third reviewer. Here, Rayyan would also used to categorize full-text screenings.

### Data Charting/Collection/Extraction

- **Data Charting Form Development:** The standard form will be developed considering JBI guidelines.
- **Data Items to be Collected:** general study characteristics (e.g., author, year, country), demographic characteristics of included populations (e.g., age, gender), participation models (e.g. active, passive, digital), determinants of participation (e.g., social, individual, systemic), outcomes (e.g., the influence of determinants on participation, the interaction between determinants), Key findings, Challenges and limitations
- **Storage of Rules for Data Extraction:** Rules of data extraction will be documented in detail and shared among all team members.
- **Data Extraction Process:** After two independent reviewers extract data based on shared rules, a pilot extraction will be done to ensure the quality. In case of disparities, the third reviewer will be consulted. If there were any missing or unclear information, we will contact the corresponding author of the included study.
- **Handling Friend Studies:** The studies conducted by the same research team, or in case of possible overlapping information, the extraction will be done carefully to avoid repeated data.

### Synthesis and Presentation of Results

- **Data Cleaning:** Data categorizing and cleaning will be performed by Microsoft excel.
- **Software for Data Cleaning and Analysis:** Microsoft Excel will also be used for qualitative analysis.
- **PRISMA Flow Diagram:** This flowchart is applied to illustrate the study inclusion and selection process.
- **Data Synthesis:** Information gathered from articles will be reported descriptively. The frequency and percentage of studies in each category, trends in publication time, and comparison among different countries will be analytically reported based on results from Microsoft Excell. We will use tables, figures, and diagrams to provide a comprehensive review of out findings.
- **Presentation of Data Items:** Data items will be reported via a comprehensive meaningful illustration to make sure findings are appliable.

### Ethics and Dissemination

- **Ethics Approval:** This scoping review, is a secondary data analysis of previously published literature does not require ethics approval.
- **Roles of Collaborators/Stakeholders:** All collaborators and stakeholders will be involved in each step of the study including developing search strategy, study selection, data extraction, and interpretation of results.
- **Dissemination Plans:** The final manuscript of this scoping review will be gone through peer-reviewed journals, conference presentations, and stakeholder meetings to widely impact decision makers.

## Conclusion

To date, there is no study comparing determinants of public participation in health care worldwide. The main goal of this scoping review is to investigate determinants of public participation worldwide to categorize the most important influencing factors of participation based on their culture, health care system, and regulations. First, we make a comprehensive picture of public participation in PHC around the world. Second, we identify the key determinants and influencing factors in addition to present challenges and future opportunities.

## Data Availability

Since this scoping review protocol does not imply the generation of new data, the review will be informed by analysis of previously published and publicly available literature. Sources of data in the review include peer-reviewed articles, gray literature, and other relevant material appropriately cited in the final review. To ensure transparency and reproducibility, the list of included studies will be made available upon publication of the review, subject to copyright and licensing restrictions of the original sources.

## Funding

This scoping review is not funded by public, commercials, or any organization.

## Conflict of Interest

The authors declare no conflicts of interest.

## Notes

### Competing Interest Statement

The authors have declared no competing interest.

### Funding Statement

This scoping review did not receive any dedicated funding from public, commercial, or not-for-profit organizations.
Conflicts of Interest
The authors declare no conflicts of interest related to this scoping review.

